# Multimorbidity and its associated risk factors among the older adults in India

**DOI:** 10.1101/2021.11.12.21265083

**Authors:** Mohd Rashid Khan, Manzoor Ahmad Malik, Saddaf Naaz Akhtar, Suryakant Yadav

## Abstract

Health at older ages is a key public health challenge especially among the developing countries. Older adults are at greater risk of vulnerability due to their physical and functional health risks. With rapidly rising ageing population and increasing burden of non-communicable diseases elderly in India are at a greater risk for multi-morbidities. Therefore, to understand this multimorbidity transition and its determinants we used a sample of older Indian adults to examine multimorbidity and its associated risk factors among the Indian elderly aged 45 and above. Using the sample of 72250 older adults this study employed the multiple regression analysis to study the risk factors of multimorbidity. Multimorbidity was computed based on the assumption of elderly having one or more than one of the diseases risks. Our results confirm the emerging diseases burden among the older adults in India. One of the significant findings of the study was the contrasting prevalence of multimorbidity among the wealthiest groups, which diverges from some earlier studies in developing countries examining the multimorbidity. Thus, given the contrasting results and rise of multimorbidity among older adults India, there is paper argues for an immediate need for proper policy measures and health system strengthening to ensure the better health of older adults in India.

**Highlights:**

- Multimorbidity is emerging as key challenge especially in the developing countries.
- There is a significant association between multimorbidity and its associated demographic and socio-economic key risk factors.
- Contrasting prevalence of multimorbidity among the affluent groups as compared to earlier studies.
- Increasing longevity has significant consequences on morbidity pattern of older adult requiring an immediate policy attention to avert the challenges of morbidity, disability and death at older ages.

## Introduction

India is witnessing an unprecedented change in the demographic and social structure in the recent decades. India is experiencing an epidemiological transition which witnesses a rising burden of noncommunicable diseases (NCDs) (Quigley 2006; Agrawal and Arokiasamy 2010, Yadav and Arokiasamy 2014). NCDs are rapidly increasing in India mainly because of lifestyle changes ^2^. With the ageing of population in India, which has now become a challenge for public health experts, policymakers, and other research organizations ^3–5^, increasing prevalence of senility is a concern in India with the rise of NCDs. There is an urgent need to understand the burden of chronic health conditions among elderly Indians, in order to improve and develop suitable responses for the future requirements of healthcare services.

Increase in longevity and decrease in mortality leads to increase the multiple comorbid conditions which is commonly known as ‘Multimorbidity’. In other words, Multimorbidity is defined as the coexistence of two or more chronic conditions which have become prevalent widely ^6,7^. Multimorbidity has now emerged as major public health issues worldwide and its associated greater adverse outcome of health like-disability, mortality, poor quality of life, hospitalizations, consequent use of medical resources and health expenditure ^8–11^.

Literatures suggests that elderly are at larger health risk due to multiple chronic diseases ^5,7,11–13^. A systematic review study has revealed that prevalence of multimorbidity among the elderly were found to be more than 55% in different countries ^14^. Despite that, high multimorbidity prevalence have been observed in many developed nations, for instance-United states, Australia ^13^, Canada ^15^ & Europe ^9,12,16^. Interestingly, the elderly from developing nations are inadequately equipped with the multimorbidity challenge, as a result a study conducted in Vietnam ^17^ revealed that more than 40% of elderly had multimorbidity conditions, whereas 69% in China ^11^ and 52% in Bangladesh ^18^. Besides that, the least developed country like Tanzania showed 25.3% multimorbidity prevalence among the elderly population.

### Multimorbidity in Indian settings

Multimorbidity research in India among the elderly is still at early stage. About 23.3% multimorbidity prevalence has been observed in India in the previous study conducted in 2017 where Kerala showed the highest prevalence of multimorbidity with 42% followed by Punjab (36%), Maharashtra (24%) & West Bengal (23%) ^19^. A recent study conducted in the district of Kerala showed 45.4% multimorbidity prevalence ^20^ Around 44% multimorbidity prevalence was found in West Bengal ^21^. A recent study conducted in Allahabad district of Uttar Pradesh showed 31% prevalence of multimorbidity ^22^.

Works of literature has suggested that there exists a strong positive association between age and the prevalence of multimorbidity in India ^19,23–25^. A study conducted in Odisha ^26^ has revealed that multimorbidity prevalence were higher among women than men and similar results have been also found in West Bengal ^21^. The rich elderly group in India were more likely to have poor health due to long term multimorbidity conditions ^27^. Recent studies have revealed that there exists significant associations between obesity ^28^ and loneliness ^29^ accompanied with multimorbidity in India. Another recent study has investigated in Odisha that multimorbidity increases the odds of elderly abuse ^30^.There are very few studies that exists on the multimorbidity prevalence and its associated risk factors among the elderly in India. Therefore, we aim to examine the prevalence of multimorbidity among the elderly in India and its states and, we examine its associated risk factors.

## Methods

### Data source

The data for this study has been taken from Longitudinal Ageing Study in India (LASI) Wave 1, which was carried out during 2017-18. LASI is a multidisciplinary, internationally harmonized panel study of 72,250 older adults aged 45 and above including their spouses less than 45 years, representative to India and all of its states and union territories (excluding Sikkim). It is a baseline data of India’s first longitudinal ageing study that provides comprehensive scientific evidence base on demographics, household economic status, chronic health conditions, symptom-based health conditions, functional health, mental health (cognition and depression), biomarkers, health insurance and healthcare utilization, family and social networks, social welfare programs, work and employment, retirement, satisfaction, and life expectations.

### Analytical sample

#### Outcome variables

The outcome variable in the study is multimorbidity which was measured on the basis of multiple chronic diseases reported among the older adults surveyed. Respondents were asked about 10 various diseases (See supplementary file) from which the outcome variable of this study was computed. These responses were combined together into trichotomous variable with categories (0= No), (1= single) and (2=more than one morbidity) to study the prevalence. But for regression analysis we converted the variable into two categories where ‘0’ represented no morbidity and ‘1’ denotes multimorbidity to apply the logistic model in the study.

#### Independent variables

Demographic and socio-economic risk factors included in the study, such as age, gender, residence, level of education, health insurance status, MPCE Quintiles, caste-group, religion, currently working, marital status (See supplemental file).

#### Statistical analysis

We used frequencies, percentages and cross tabulations for prevalence of mmultimorbidity with respect to the social and demographic characteristics with 95% confidence interval. We applied chi-square test (χ2) to see the association between multimorbidity and its covariates. We then performed the logistic regression to study the determinants of multimorbidity among older adults in India

## Results

### Socio-demographic characteristics

The socio-economic and demographic characteristics of the study population by the number of diseases are presented in **Table 1**. The mean number of illnesses per year caused by single disease in the sample was 2.58 (SD = 2.19) with 18.75% (95% CI 18.16% - 19.36%), whereas it was 62.68% (95% CI 61.9% - 63.45%) for more than one disease, i.e., multimorbidity.

**Table 1.** Mean score of morbidity among the older-adults in India (N=72250).

| Indicators | Mean Score of morbidity |
| --- | --- |
|  | Mean (SD) |
| Overall | 2.58 (2.19) |
| Multimorbidity - No | 0.50 (0.50) |
| Multimorbidity - Yes | 3.79 (1.86) |

*Source: Authors' calculation using LASI-wave-1 data*

### Multimorbidity prevalence at national level

Table 2 shows the prevalence of multimorbidity increased substantially with age, from 44.69% (95% CI = 39.3% - 50.22%) in ≥ 40years to 74.12% (95% CI = 70.43% - 77.5%) in those aged above 80 years. Similarly, the crude prevalence of multimorbidity increased modestly with increasing household wealth, from 53.78% (95% CI = 52.32% - 55.23%) in the lowest wealth quintile to 71.97% (95% CI = 70.02% - 73.84%) in the highest wealth quintile. There is considerably higher prevalence of multimorbidity in urban 69.59% (95% CI = 67.66% - 71.45%) against rural area 59.48% (95% CI = 58.79% - 60.17%). There seems not much difference in male and female as far as multimorbidity is concern, however, widowed has higher prevalence of multimorbidity 69.51% (95% CI = 67.98% - 71.00%) compare to currently married 60.92% (95% CI = 60.04% - 61.78%).

**Table 2.**
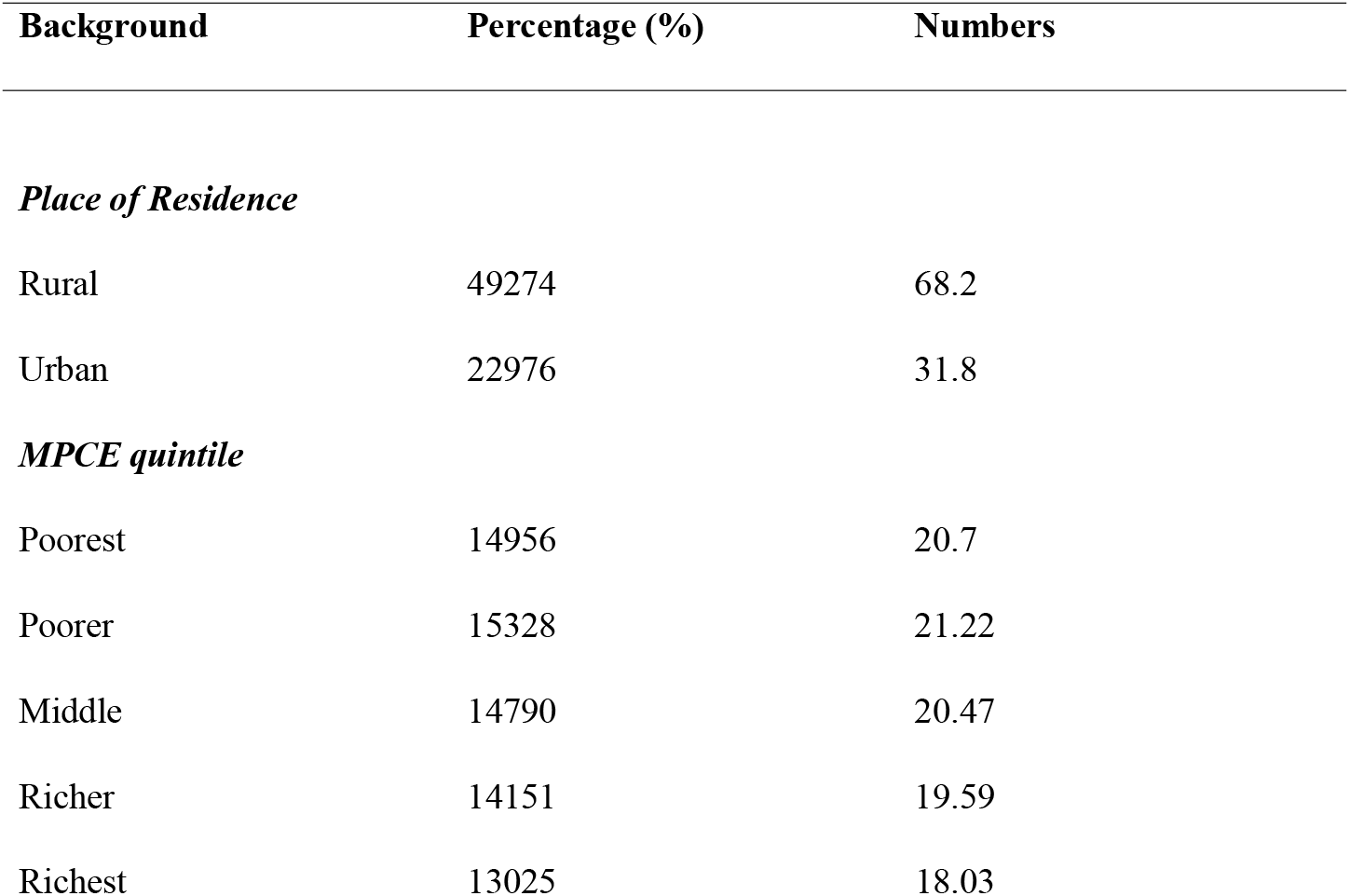
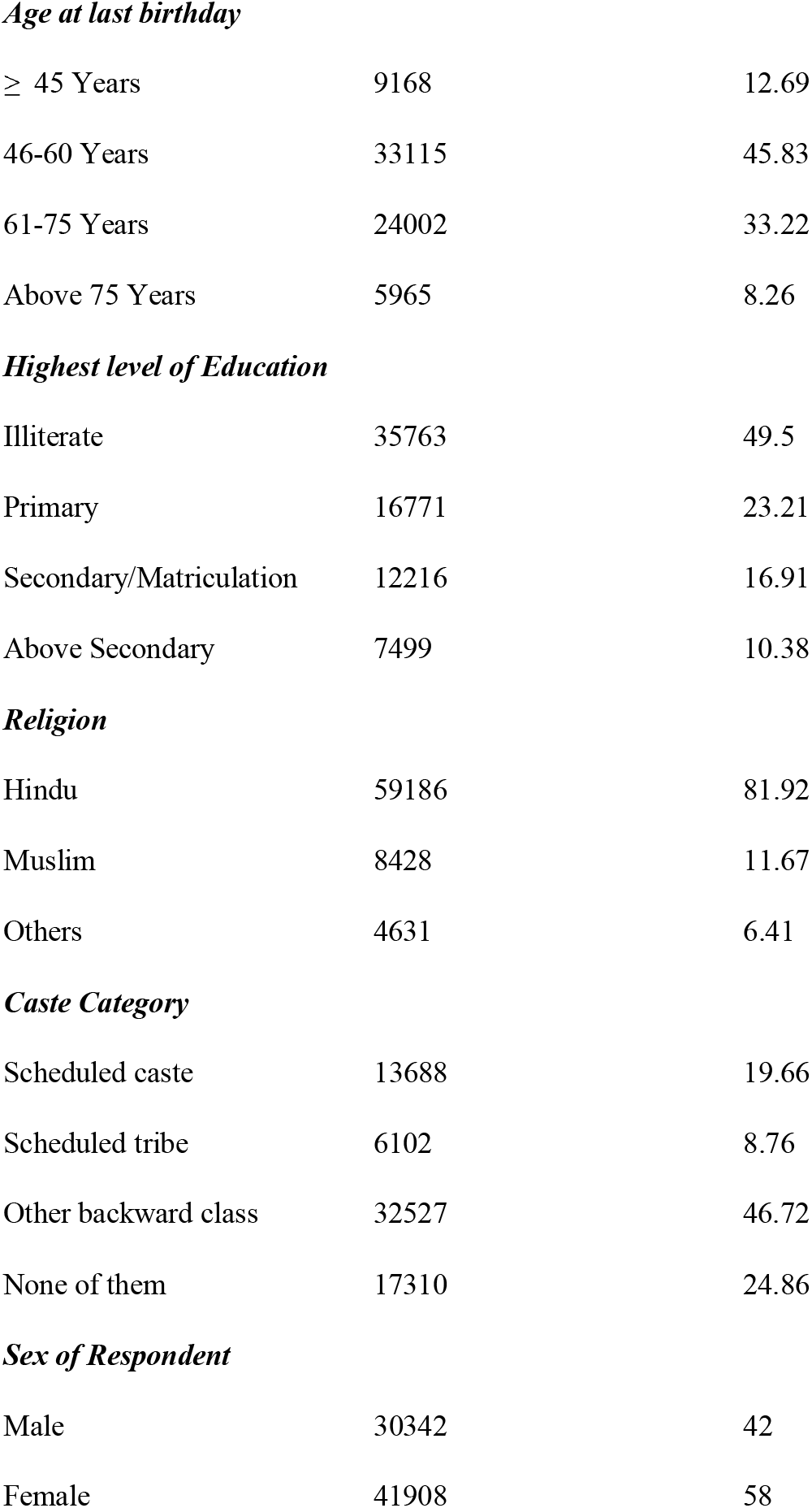
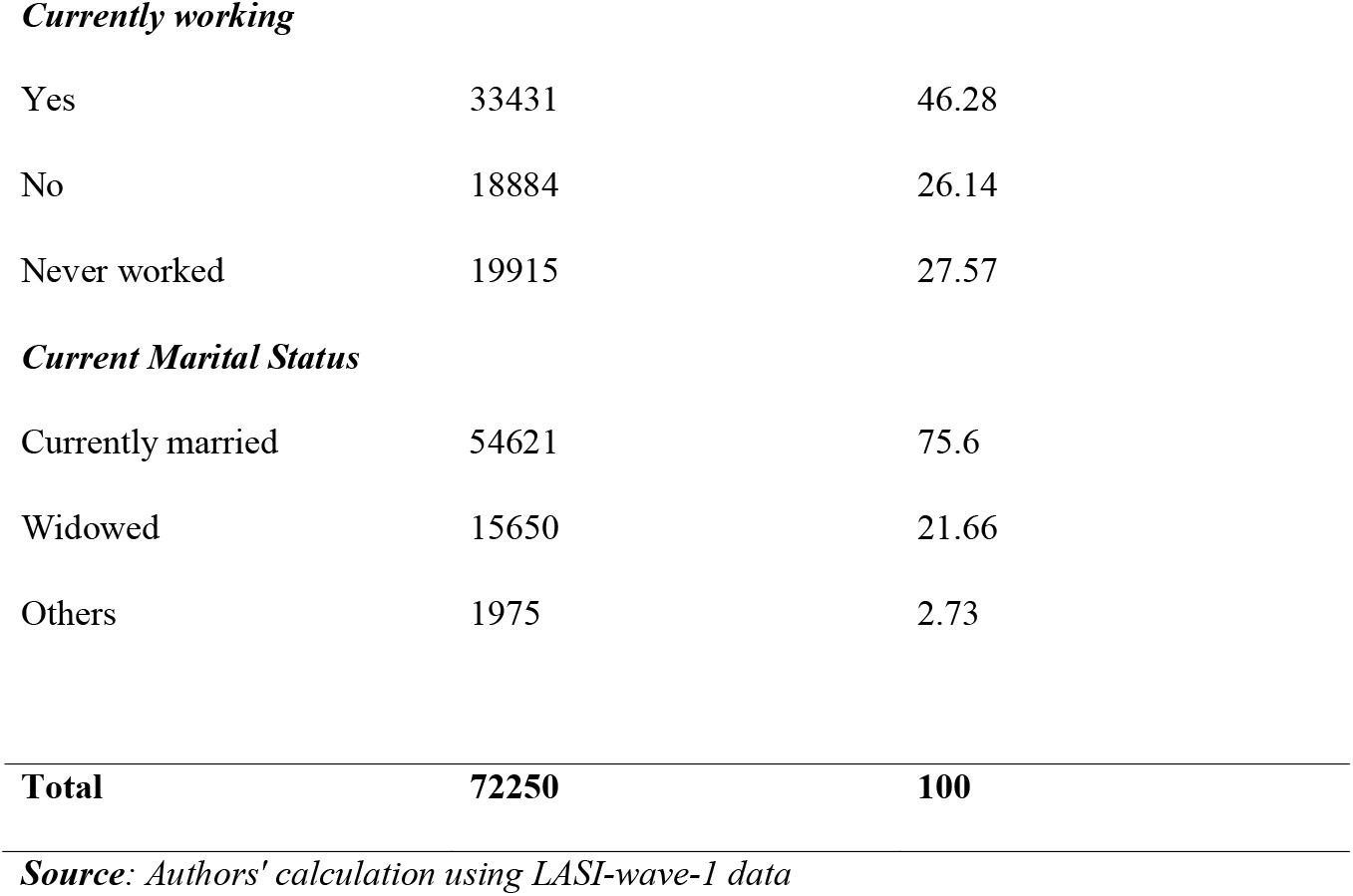
Sample distribution showing morbidity among the older-adults in India with suitable socio-demographic and economic background (N=72250).

| Background | Percentage (%) | Numbers |
| --- | --- | --- |
| *Place of Residence* |  |  |
| Rural | 49274 | 68.2 |
| Urban | 22976 | 31.8 |
| *MPCE quintile* |  |  |
| Poorest | 14956 | 20.7 |
| Poorer | 15328 | 21.22 |
| Middle | 14790 | 20.47 |
| Richer | 14151 | 19.59 |
| Richest | 13025 | 18.03 |
| ***Age at last birthday*** |  |  |
| ≥ 45 Years | 9168 | 12.69 |
| 46-60 Years | 33115 | 45.83 |
| 61-75 Years | 24002 | 33.22 |
| Above 75 Years | 5965 | 8.26 |
| ***Highest level of Education*** |  |  |
| Illiterate | 35763 | 49.5 |
| Primary | 16771 | 23.21 |
| Secondary/Matriculation | 12216 | 16.91 |
| Above Secondary | 7499 | 10.38 |
| ***Religion*** |  |  |
| Hindu | 59186 | 81.92 |
| Muslim | 8428 | 11.67 |
| Others | 4631 | 6.41 |
| ***Caste Category*** |  |  |
| Scheduled caste | 13688 | 19.66 |
| Scheduled tribe | 6102 | 8.76 |
| Other backward class | 32527 | 46.72 |
| None of them | 17310 | 24.86 |
| ***Sex of Respondent*** |  |  |
| Male | 30342 | 42 |
| Female | 41908 | 58 |
| ***Currently working*** |  |  |
| Yes | 33431 | 46.28 |
| No | 18884 | 26.14 |
| Never worked | 19915 | 27.57 |
| ***Current Marital Status*** |  |  |
| Currently married | 54621 | 75.6 |
| Widowed | 15650 | 21.66 |
| Others | 1975 | 2.73 |
| **Total** | **72250** | **100** |

*Source: Authors' calculation using LASI-wave-1 data*

**Table 3.**
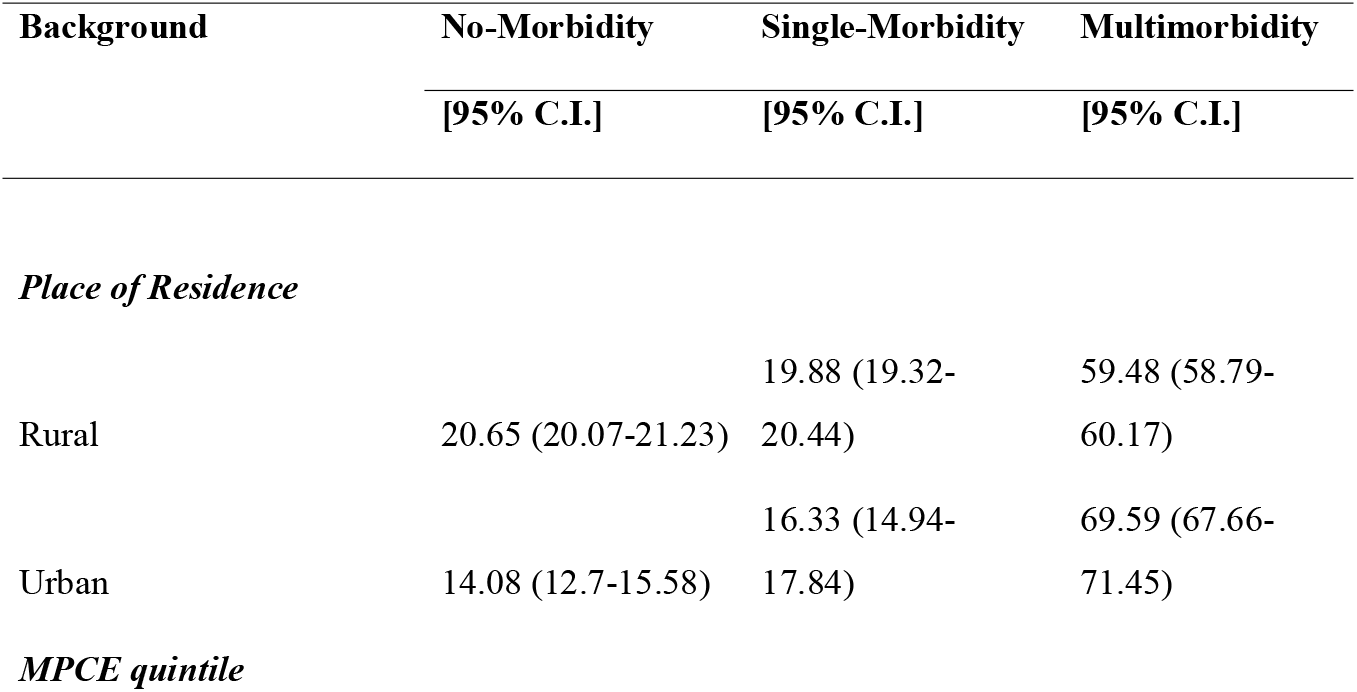
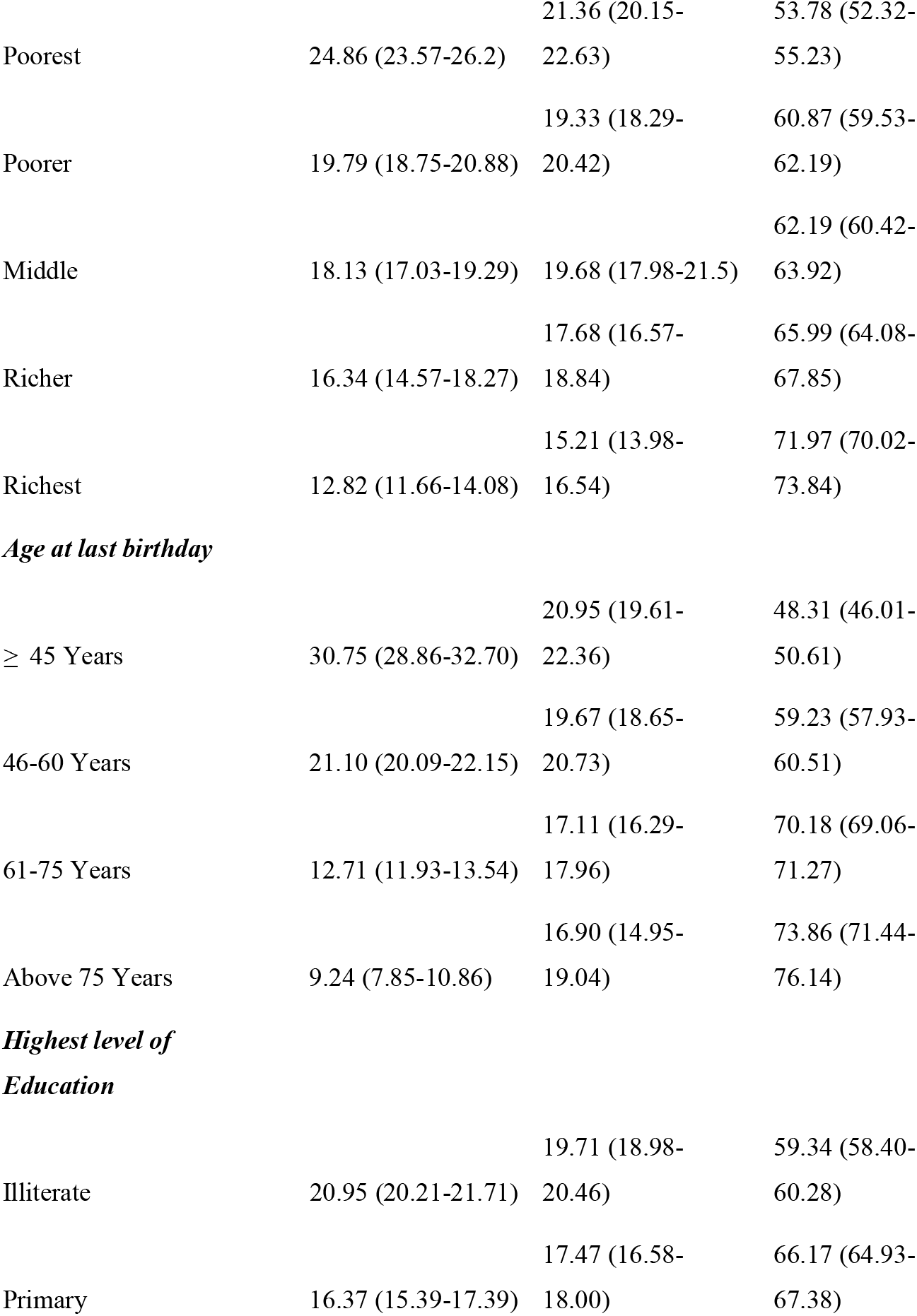
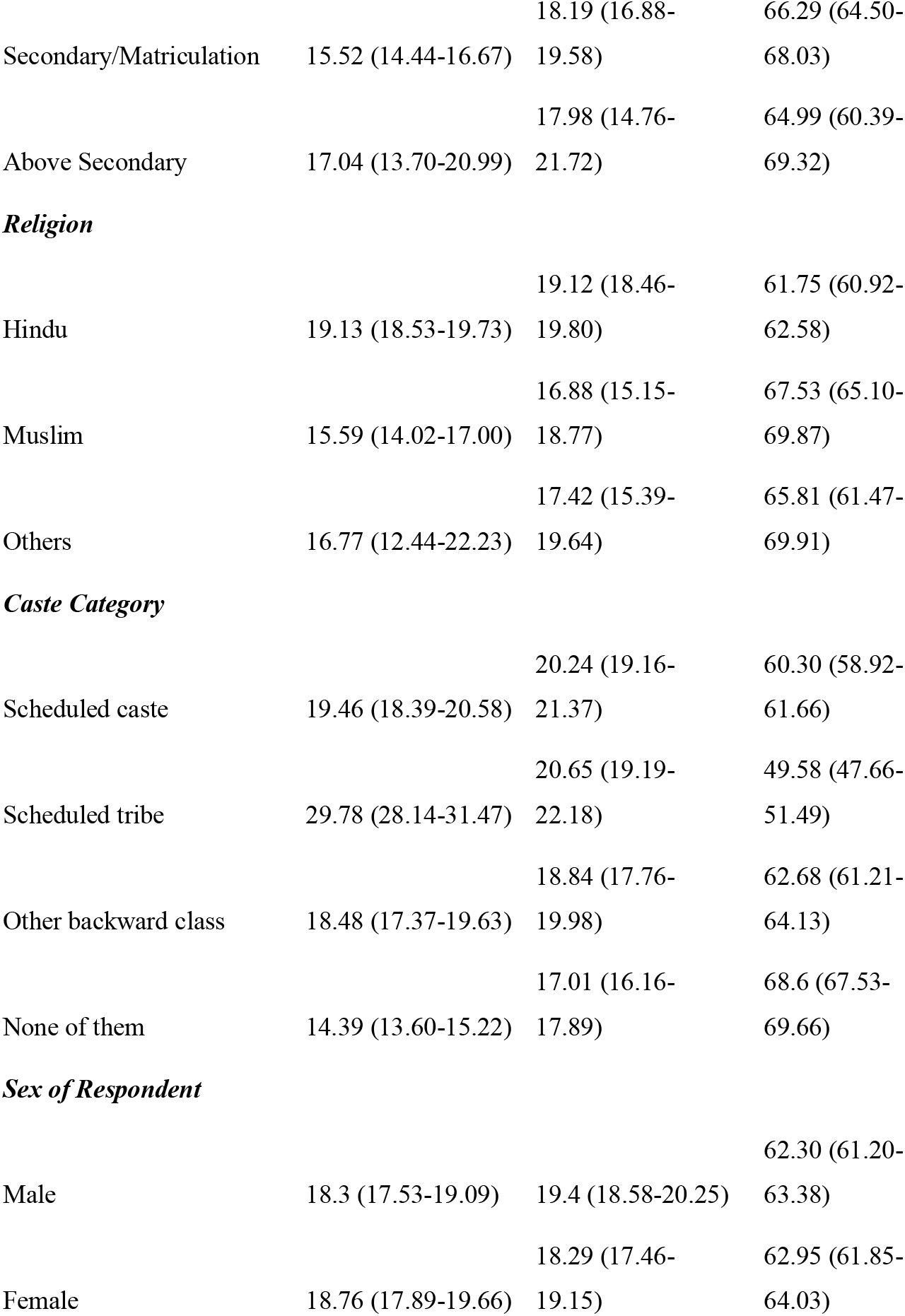
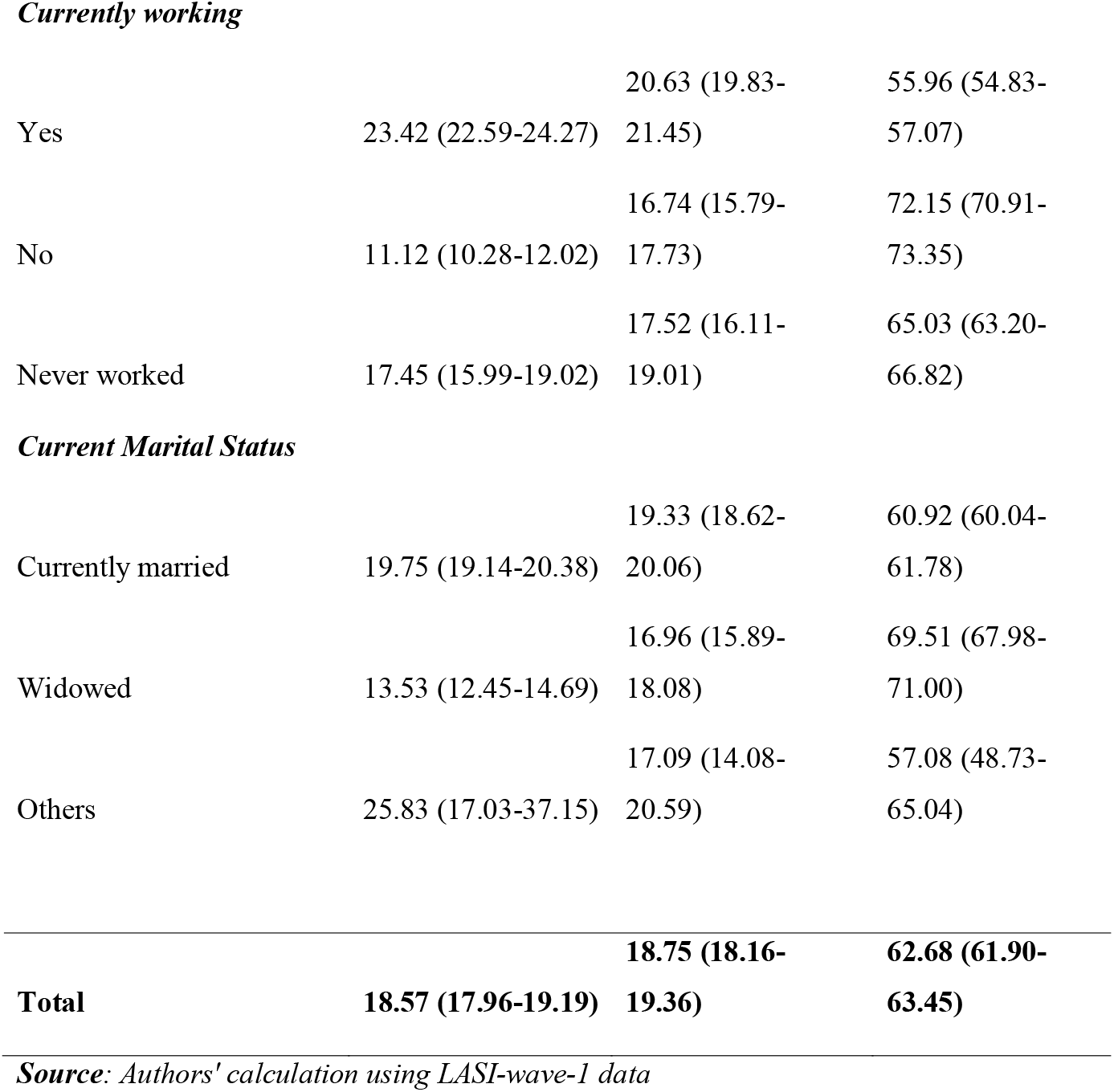
Prevalence of morbidity (%) among the older-adults in India with suitable socio-demographic and economic background (N=72250).

**Table 4.**
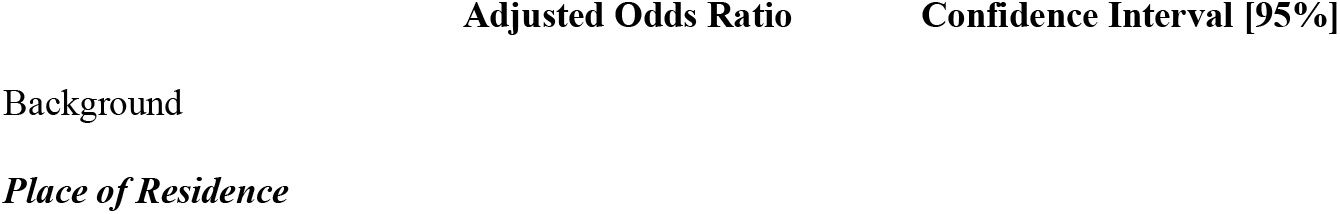
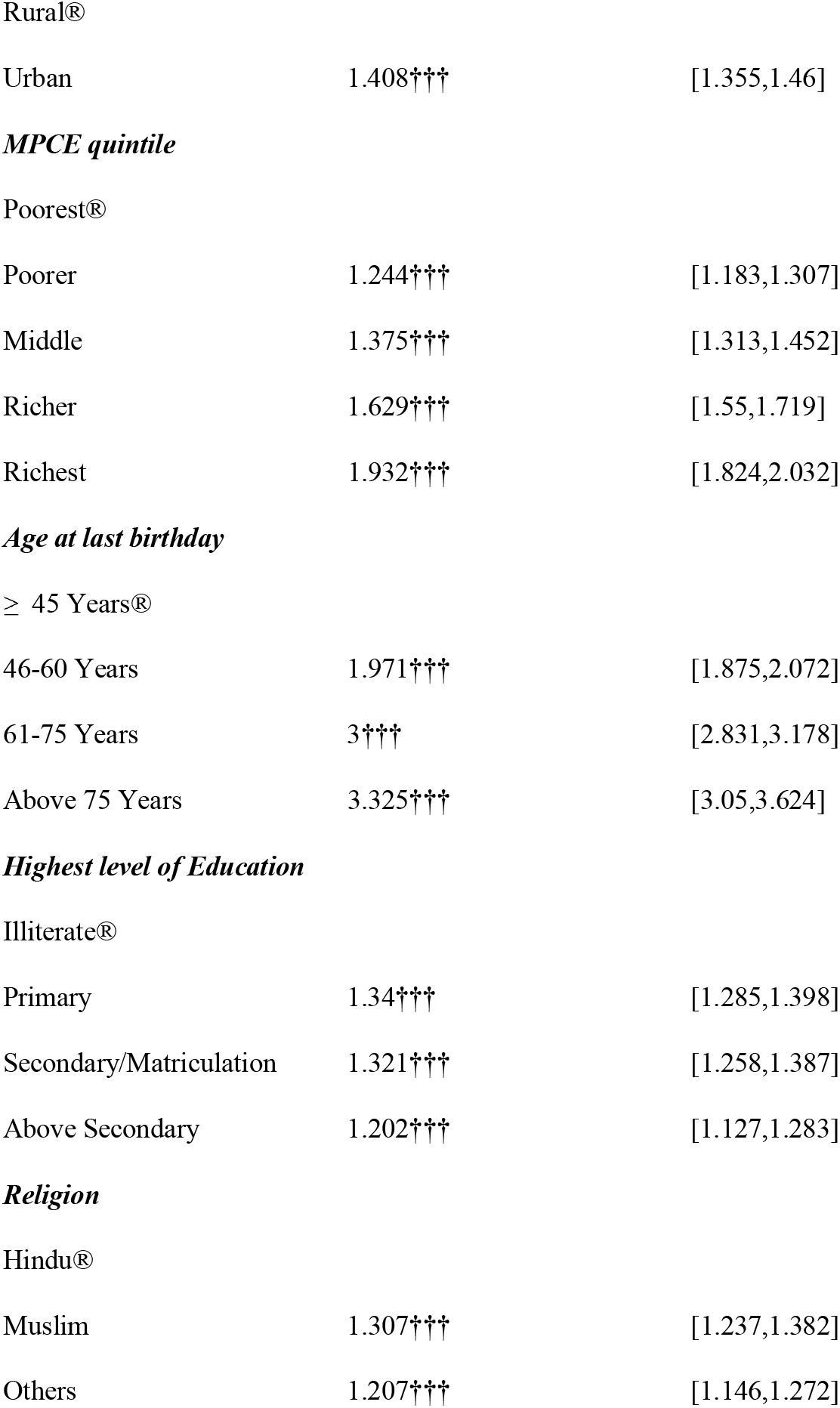
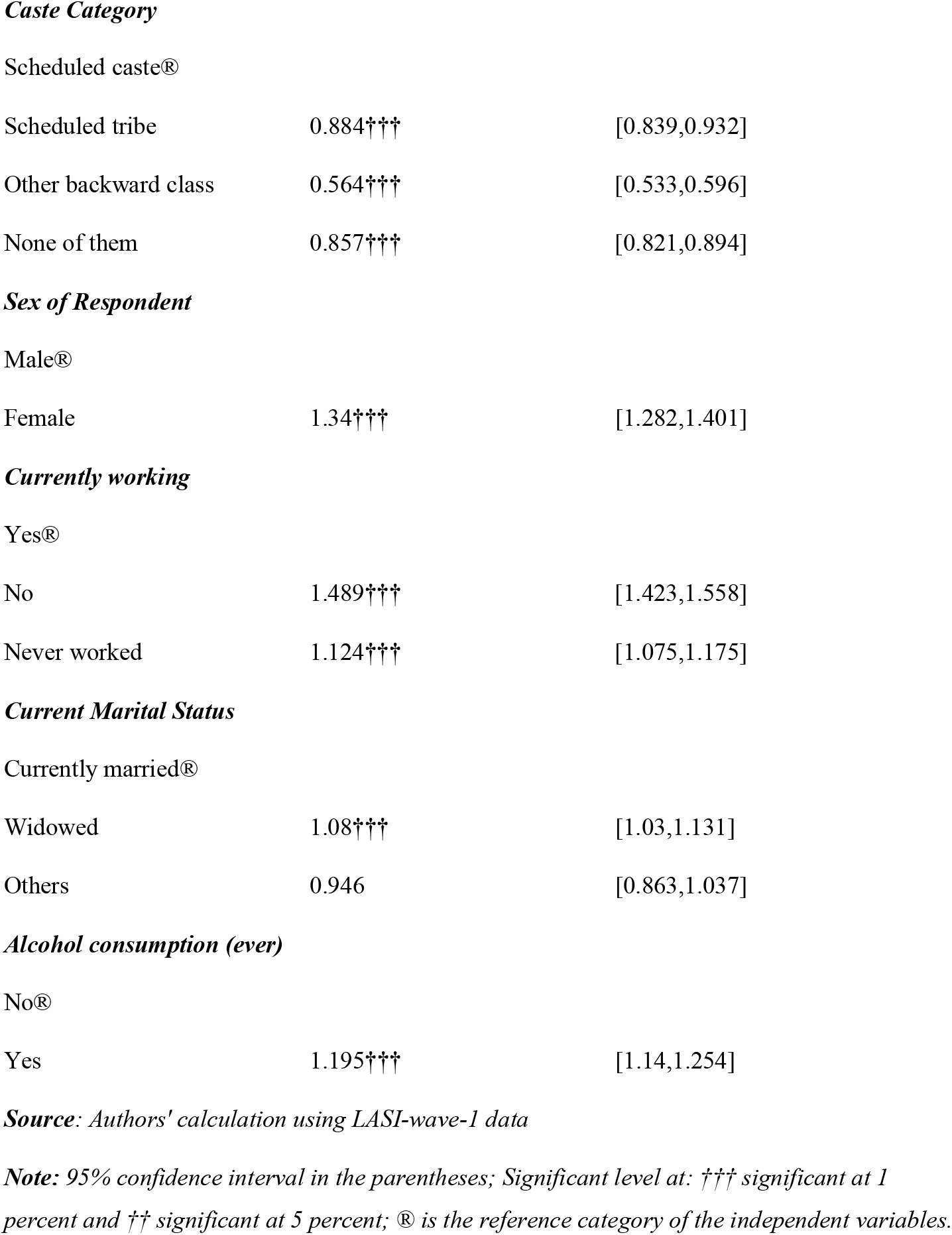
Regression analysis result showing the associated risk factors of multimorbidity among the older-adults in India (N=72250).

|  | **Adjusted Odds Ratio** | **Confidence Interval [95%]** |
| --- | --- | --- |
| Background |  |  |
| *Place of Residence* |  |  |
| Rural® |  |  |
| Urban | 1.408††† | [1.355,1.46] |
| Poorer | 1.244††† | [1.183,1.307] |
| Middle | 1.375††† | [1.313,1.452] |
| Richer | 1.629††† | [1.55,1.719] |
| Richest | 1.932††† | [1.824,2.032] |
| 46-60 Years | 1.971††† | [1.875,2.072] |
| 61-75 Years | 3††† | [2.831,3.178] |
| Above 75 Years | 3.325††† | [3.05,3.624] |
| Primary | 1.34††† | [1.285,1.398] |
| Secondary/Matriculation | 1.321††† | [1.258,1.387] |
| Above Secondary | 1.202††† | [1.127,1.283] |
| Muslim | 1.307††† | [1.237,1.382] |
| Others | 1.207††† | [1.146,1.272] |
| Scheduled tribe | 0.884††† | [0.839,0.932] |
| Other backward class | 0.564††† | [0.533,0.596] |
| None of them | 0.857††† | [0.821,0.894] |
| Female | 1.34††† | [1.282,1.401] |
| No | 1.489††† | [1.423,1.558] |
| Never worked | 1.124††† | [1.075,1.175] |
| Widowed | 1.08††† | [1.03,1.131] |
| Others | 0.946 | [0.863,1.037] |
| Yes | 1.195††† | [1.14,1.254] |

***Source:*** Authors' calculation using *LASI-wave-1* data***Note:*** 95% confidence interval in the parentheses; Significant level at: ††† significant at 1 percent and †† significant at 5 percent; ® is the reference category of the independent variables.

### Single & multimorbidity prevalence at state level

**Map 1** shows the single morbidity prevalence among the elderly in India at state level using LASI Wave-1 data. The highest single morbidity prevalence was in Odisha (24.4%), followed by Assam (23.2%), Chhattisgarh (23%) & Tamil Nadu (22%) whereas the lowest were seen in Punjab (10.9%), and followed by Kerala (13.4%), Meghalaya (14%) & Chandigarh (14%). **Map 2** indicates the multimorbidity prevalence among the elderly in India at state level. The prevalence of multimorbidity was the highest in Punjab (83%), Chandigarh (78.7%), Kerala (78%), West Bengal (73.4%), & Goa (72.5%) while the lowest were found in Nagaland with 42.6%, followed by Chhattisgarh (44.6%), Meghalaya (48.8%), Odisha (49.4%) & Jharkhand (51.5%).

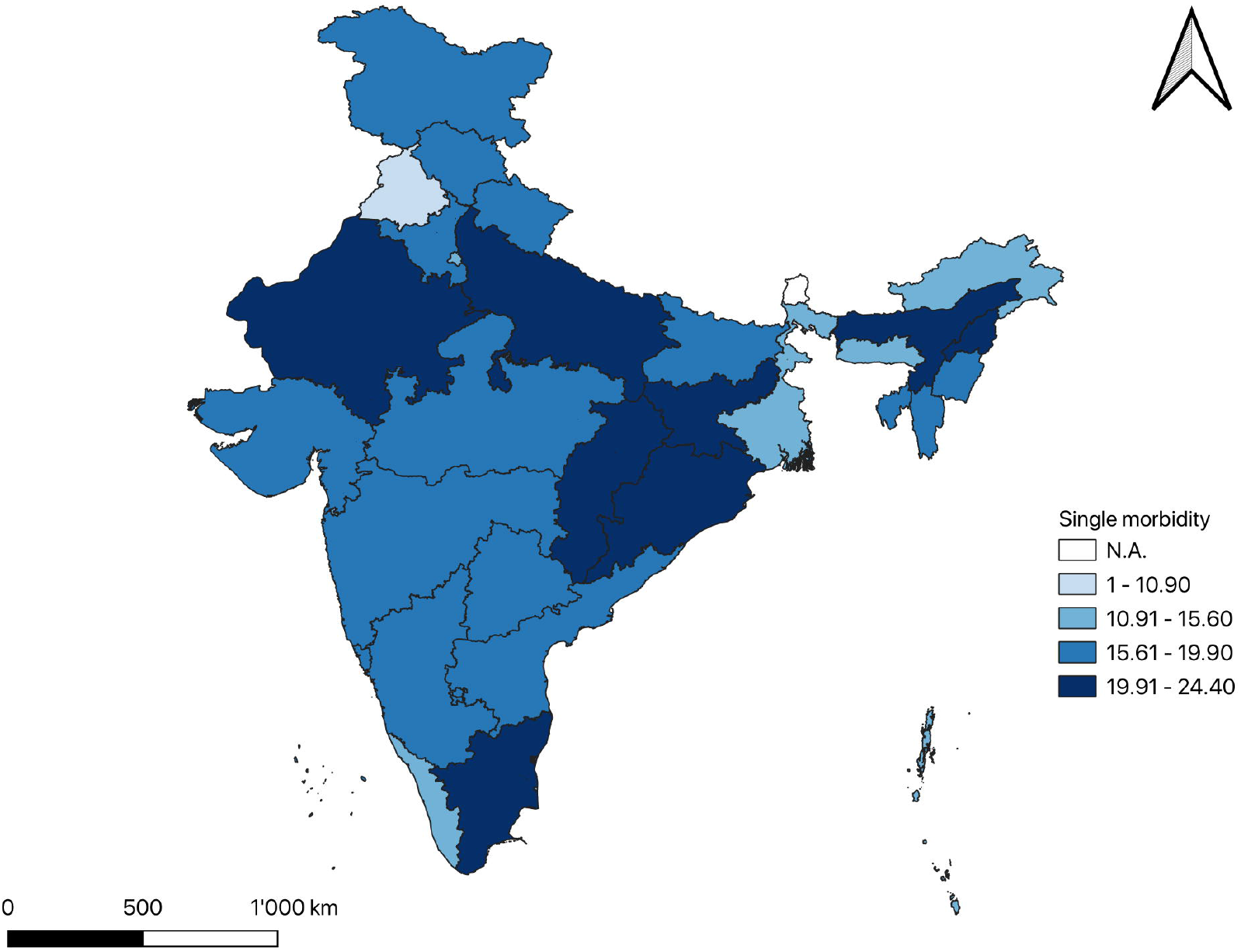

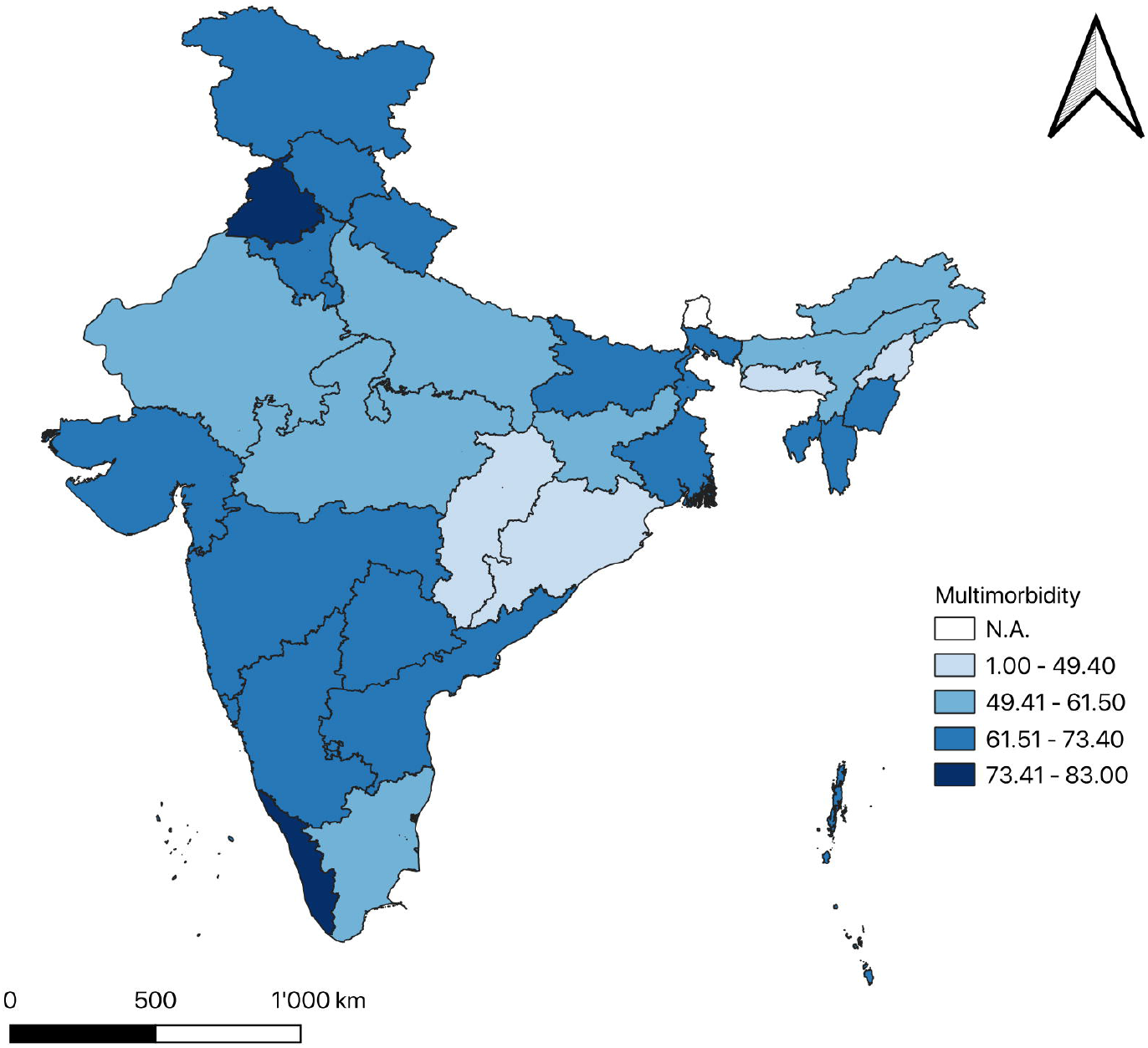

### Determinants of Multimorbidity

Multimorbidity is common at higher ages given the associated risk of physical and functional vulnerability. There are numerous other risk factors like smoking drinking, underweight obesity, physical limitations and occupational exposures that are likely to enhance the risk for multimorbidity. Multivariate associations between population characteristics and multimorbidity are presented in Table 2. The predicted probability of having a multiple disease showed a significant increase, and it is almost 5 times more likely in 71-80 years (Adjusted OR=5.033; 95% CI = 4.553 - 5.564; reference category ≥ 40 years of age). Women were more likely than men to have more than one morbidity (Adjusted OR=1.354; 95% CI = 1.296 - 1.416). Other characteristics like urban resident (OR= 1.406; 95% CI = 1.355 - 1.460) with reference to rural, Muslim (AOR= 1.322; 95% CI = 1.250 - 1.398) against Hindu, currently working (AOR= 1.436; 95% CI = 1.373 - 1.503), ever consumed any alcoholic beverages (AOR= 1.198; 95% CI = 1.142 - 1.257) and widowed (AOR= 1.062; 95% CI = 1.014 - 1.113) against currently married have higher likelihood of having multimorbidity. The lower caste categories SC (AOR= 0.891; 95% CI = 0.845 - 0.940), ST (AOR= 0.568; 95% CI = 0.537 - 0.601) and OBC (AOR= 0.861; 95% CI = 0.825 - 0.899) are less likely of having multimorbidity compared with the people in Other (Upper Hindu caste) category.

## Discussion

Multimorbidity is emerging as a critical public health challenges, especially, in the developing countries such as India. Owing to the life style changes and shift in disease pattern and rise in out-of-pocket expenditure (OOPE), multimorbidity is resulting into an economic burden for countries. In parallel to the rise in multimorbidity, ageing of population with increase in life expectancy has become a major public health challenge. The ageing of population further manifests the multifold vulnerability in old ages caused by these diseases’ risks. In the view to rise in risk of diseases, we examined the prevalence and risk factors of multimorbidity in 45 and above years using data provided in the LASI wave-1 in India.

An individual suffers from multimorbidity due to multiple reasons ranging from comorbidities that may arise due to a common risk factor or due to the outcome of a particular diseases leading to other diseases ^31^. This risk likely enhances with age due to physical and functional vulnerabilities. Research shows ageing contributes to multimorbidity through the loss of physical and functional health including frailty, which later results into greater complications like falls, disability, immobility, and mortality ^32,33^.

Our results showed the significant association between multimorbidity and its associated demographic and socio-economic risk factors like age, income, education and place of residence. The results corroborate with the earlier findings where significant association was found between multimorbidity and socio-economic outcomes ^34^.

This study showed the significant and positive relation of multimorbidity in urban areas. The risk associated with multimorbidity is higher in 45 and above years in urban areas as compared to rural areas. This higher risk in urban areas is likely attributable to increasing life style changes ^35^. This higher risk of disease in urban areas is also appreciated due to the imbalance in medical care that exists in weak health care facilities ^36^.

One of the significant findings of this paper is the contrasting prevalence of multimorbidity among the most wealthiest groups, which diverges from some earlier studies from developing countries examining the multimorbidity ^37,38^. One of the most likely reasons maybe self-reporting of morbidity given the fact that elderly belonging to better socio-economic classes have greater access to health care service provisions, which increases the likelihood of their diagnosis and care for a particular diseases ^39^.

Multimorbidity increases likely to ageing risk as shown by various studies ^33,40^. These findings are also well reflected through our results where increase in age likely enhances the risk for more than one morbidity. Therefore, increasing longevity has a likely consequences of morbidity pattern of older adults, which needs an immediate policy attention to avert the challenges of morbidity, disability and death at older ages. Furthermore, strong measures can ensure the active and healthy ageing interventions to avert the diseases burden with greater concentration of older adults in upper ages.

## Conclusions

This study provides an evidence of emerging diseases burden among the older adults in India. The study highlights the need for better interventions for older adults to avert the health crisis in later years of life. As the findings of this research specifically indicate the growing burden of multimorbidity, there is an immediate need for proper policy measures and health system strengthening to ensure the health ageing in India. Moreover emphasis should be given on workforce training and quality improvement strategies which can ensure the better physical and functional health of older adults. There is also an immediate need for improving the financial incentives for elderly at older ages given the challenges they face in terms of health and social security provisions in India.

## Data Availability

The LASI Wave-1 data was collected by the nodal institution International Institute for Population Sciences, Mumbai on behalf of the Ministry of Health and Family Welfare, Government of India. All data were de-identified. The de-identified version of the LASI Wave-1 data is publicly available to the researchers and policymakers upon formal request to the International Institute For Population Sciences for access (link to the data request document LASI_DataRequestForm_0.pdf (http://www.iipsindia.ac.in) and link for the other information for lasi data set LASI Wave-I | International Institute for Population Sciences (IIPS) (http://www.iipsindia.ac.in).

## Acknowledgements

No acknowledgements

