## Supplemental 1 for "Multimorbidity and its associated risk factors among the older adults in India": Supplemental file.docx

**Respondent information:**

The main variable of interest is whether the respondent had more than one disease. We defined respondents as having an illness if they answered affirmatively the following two questions: “Have you ever been told by a health professional that you have.? (For example, Diabetes or high blood sugar)”, or “Have you ever been diagnosed with?”. We counted the number of health conditions for each respondent, and defined those with multimorbidity as the presence of two or more of the above listed conditions without a specific reference condition.

**The list of diseases used in the study:**

1. **Non communicable diseases:** Hypertension or high blood pressure; Diabetes or high blood sugar; Cancer or a malignant tumor; Chronic lung disease such as asthma ,chronic obstructive pulmonary disease/Chronic bronchitis or other chronic lung problems; Chronic heart diseases such as Coronary heart disease (heart attack or Myocardial Infarction), congestive heart failure, or other chronic heart problems; Stroke; Arthritis or rheumatism; Osteoporosis or other bone/joint diseases; Any neurological, or psychiatric problems such as depression, Alzheimer’s/Dementia, unipolar/bipolar disorders, convulsions, Parkinson’s etc.; High cholesterol.
2. **Other NCDs:** Thyroid disorder, Gastrointestinal problems (GERD, constipation, indigestion, piles, peptic Ulcer), Skin diseases and others.
3. **Urogenital:** Chronic Renal Failure, Incontinence, Kidney Stones, BPH (Benign Prostatic Hyperplasia).
4. **Eyesight:** Presbyopia, Cataract, Glaucoma, Myopia (Nearsightedness), Hypermetropia (Farsightedness).
5. **Ear-related problem:**
6. **Oral health conditions:** Painful teeth, Ulcers lasting more than two weeks, bleeding gums, swelling gums, welling gums, dental cavity/dental caries, Soreness or cracks in the corner of the mouth and others.

**Description of independent variables**

**Age at last birthday:** Below 45 years, 46-60 years, 61-75 years, Above 75 years;

**Gender:** Male and Female;

**Residence:** Rural and Urban

**Level of Education:** No formal education, Primary school completed, Secondary/matriculation and above secondary;

**Health Insurance Status:** With insurance and without insurance;

**MPCE Quintiles:** Poorest, Poorer, Middle, Richer and Richest;

**Caste Category:** Scheduled caste, Scheduled tribe, Other backward class and None of them; **Religion:** Hindu, Muslim and Others;

**Currently working:** No, Yes and never worked;

**Current marital status:** Currently married, Widowed, others.

**Table S1** Showing the prevalence (%) of single morbidity and multimorbidity in 35 Indian states.

| **State** | **Single** | **Multimorbidity** |
| --- | --- | --- |
| Andaman & Nicobar Island | 14.34 | 72.11 |
| Andhra Pradesh | 18.35 | 68.47 |
| Arunachal Pradesh | 14.34 | 54.98 |
| Assam | 23.24 | 53.83 |
| Bihar | 17.42 | 64.81 |
| Chandigarh | 14.02 | 78.67 |
| Chhattisgarh | 22.97 | 44.62 |
| Dadra & Nagar Haveli | 14.47 | 68.76 |
| Daman & Diu | 18.53 | 69.65 |
| Delhi | 15.6 | 70.21 |
| Goa | 17.11 | 72.52 |
| Gujarat | 17.36 | 65.18 |
| Haryana | 18.23 | 71.37 |
| Himachal Pradesh | 17.66 | 70.86 |
| Jammu & Kashmir | 19.68 | 66.21 |
| Jharkhand | 21.21 | 51.47 |
| Karnataka | 18.32 | 63.79 |
| Kerala | 13.37 | 77.99 |
| Lakshadweep | 19.04 | 70.2 |
| Madhya Pradesh | 18.43 | 54.5 |
| Maharashtra | 17.85 | 65.4 |
| Manipur | 18.88 | 69.66 |
| Meghalaya | 13.95 | 48.83 |
| Mizoram | 18.84 | 67.66 |
| Nagaland | 21.91 | 42.6 |
| Odisha | 24.39 | 49.4 |
| Puducherry | 15.43 | 68.14 |
| Punjab | 10.93 | 82.95 |
| Rajasthan | 21.55 | 55.49 |
| Tamil Nadu | 22.04 | 61.47 |
| Telangana | 19.94 | 65.65 |
| Tripura | 17.84 | 67.46 |
| Uttar Pradesh | 20.87 | 55.55 |
| Uttarakhand | 17.79 | 68.76 |
| West Bengal | 14.89 | 73.38 |
